# The spectrum of rare and novel indel mutations responsible for β Thalassemia in eastern India

**DOI:** 10.1101/2022.08.29.22279342

**Authors:** Sajan Sinha, Paramita Bhattacharya, Mrinal Kanti Das, Atanu Kumar Dutta

## Abstract

**Purpose:** There is limited data available regarding the clinical utility of routine molecular diagnosis of β Thalassaemia in addition to HPLC-based screening in low resource settings. The current study highlights the caveats of an HPLC-based screening compared to the inclusion of genetic confirmation as a second-tier test and its implications in terms of genotype-phenotype correlation.

**Material and Methods:** A prospective, institution-based, observational study was conducted at the Department of Paediatric Medicine, including 103 children aged up to 12 years. Five common mutations for β Thalassemia and the HbE mutation in the *HBB* gene were tested by a two-tiered approach using multiplex ARMS PCR and PCR RFLP methods respectively. Sanger sequencing of all three exons of the *HBB* gene was performed in all negative cases.

**Results:** Sequencing revealed many rare pathogenic mutations like c.316-106C>G (dbSNP: 34690599); Hb Kairouan (c.92G>C); c.33C>A (dbSNP rs35799536); c.47G>A (dbSNP rs63750783); c.51delC (HbVar ID 799); c.[93-2A>C] and c.118C>T (HbVar ID 845). We detected a novel Pathogenic M_000518.5(*HBB*):c.164_168delinsGGCATCA (p.Val55fs) mutation in a heterozygous state which was reported in the ClinVar database with accession ID VCV000590977.2. We also encountered several cases of silent carrier on HPLC and de novo occurrence of mutation.

**Conclusion:** We conclude that the multiplex touchdown ARMS PCR methodology employed in the present study provides a low-cost solution for molecular diagnostics of Β Thalassaemia. The problem of silent carriers in HPLC is significant enough to rethink if we need supplemental genetic testing in the couple when one of the partners is a carrier.

## Introduction

India has a huge burden with an estimated 100,000 patients with β Thalassemia syndrome and every year adding 8,000 to 10,000 [1]. Hemoglobin HPLC is well established as confirmatory investigation for the disease but the problem of silent carriers needs supplemental genetic testing in the couple when one partner is carrier. More over if the patient presents very early within first year of life in remote areas and starts getting blood transfusion every month then Hb HPLC study for the diagnosis of Thalassemia syndrome is not conclusive. Therefore, genetic mutation study has got definite role in this group of patients for the confirmation of the diagnosis of the disease. Out of the multitude of mutations commonest five are: 1. IVS 1-5 - NM_000518.4:c.92+5G>C, rs33915217, 2. 619 base del -NG_000007.3:g.71609-72227del619, 3. IVS 1-1 - NM_000518.4:c.92+1 G>A, rs33971440, 4. CD 8/9 - NM_000518.4:c.27dupG rs35699606, 5. CD 41/42 - NM_000518.4:c.126-129delCTTT, rs281864900. Number of studies have highlighted the burden of β Thalassaemia in India and the mutation spectrum [1-5]. Study done by Bhattacharyya DM *et al* on 660 individuals in West Bengal revealed the prevalence of IVS1-5 mutation among the studied β carriers to be 46.6 %, and codon 26 (G>A) mutation to be 31.54 % [6]. Other prevailing mutations among the screened individuals include codon 30 (7.53 %), codon 15 (5.01 %), codon 41/42 (3.58 %), and codon 8/9 (1.07 %). There were several studies showing discordant results on genotype phenotype correlation [7-11]. Among the genetic modifiers some prominent factors are alpha haemoglobin gene triplication [8], XmnI polymorphism [9 and]concurrent alpha thalassemia carrier status [10].

There is lack of studies in β Thalassemia involving cost effective yet comprehensive mutation detection methods. The current study was envisaged to fill the gap of knowledge in this field along with highlighting some other relevant observations. Therefore, a prospective, institution based, observational study was conducted including 103 children aged up to 12 years, who were already diagnosed or were diagnosed during the study period as cases of β Thalassemia, meeting the inclusion and exclusion criteria. The aim of the study was to find the mutation spectrum, establish genotype phenotype correlation and assess the utility of routine genetic testing vs a second-tier approach to genetic testing combined with hemoglobin HPLC. In this study we have employed a modified version of the multiplex ARMS PCR developed by Bhardwaj U *et al* [12] which enabled us to investigate the five common mutation at a cost of less than 2 USD per subject.

## Materials and Methods

A hospital based, prospective, observational study was carried out in the inpatient and outpatient department of Paediatric Medicine and Thalassemia Control Unit of a tertiary care centre in West Bengal, Eastern India. Genetic testing was carried out at the same hospital. In this study, a population of 103 patients with symptomatic β Thalassemia were evaluated. All the children attended the study place in presence of the principal investigator satisfying the inclusion and exclusion criteria during the study period were included in the study till the target sample size was reached.

### Inclusion criteria

1. Age up to 12 yrs.
2. Children with chronic haemolytic anaemia diagnosed by HPLC of hemoglobin as patient of β Thalassemia syndrome.
3. Chronic haemolytic anaemia but hemoglobin HPLC was inconclusive for the β Thalassemia diagnosis which was subsequently confirmed by genetic mutation study.
4. Available adequate medical records supplemented with history from reliable sources.
5. Parents or legally acceptable / authorized representative (LAR) of patient who were agreed for giving written informed consent as well as children aged 7 to 12 years who were giving verbal assent in addition.

### Exclusion criteria

1. Chronic haemolytic anaemia other than β Thalassemia.
2. Received packed red blood cell transfusion within last 2 weeks.
3. Hemoglobin HPLC inconclusive or not feasible and subsequently Genetic mutation study did not detect any mutation.

Data was collected by filling the pre-structured questionnaire proforma through relevant history taking and data collecting from previous medical records. History including clinic-epidemiological information’s and detailed treatment and relevant laboratory records of the patients were studied in the light of the following parameters: age and sex; age at first blood transfusion; age at diagnosis; frequency of blood transfusions per year; mean haemoglobin at the time of diagnosis; haemoglobin HPLC results; genetic Study(ARMS PCR, and Sanger sequencing. After obtaining ethical clearance from the Institutional Ethics Committee, study was conducted among the study population after taking written informed consent / assent from the legal guardians/patients respectively. Hemoglobin HPLC study when required was done in Thalassemia control unit of this institute. Based on the age of first presentation, HPLC reports and total number of transfusions study population were grouped as TDT (Transfusion Dependent Thalassemia) and NTDT (Non -Transfusion Dependent Thalassemia).

### Genotyping

From all patients 2 (two) ml blood was collected in EDTA tube and stored in −200C freezer until DNA extraction. DNA was extracted by salting out method. Common five β Thalassemia mutations were detected by a previously described multiplex ARMS-PCR method and if required by sanger sequencing. Then the data collected through the study was analysed accordingly. The detailed genotyping methodology is presented in the supplementary material.

### Statistical data analysis

Data was collected, recorded and compiled on Microsoft Excel data sheet, statistical methods (mean, standard deviation) and software’s were used to analyse the data. Frequencies of various signs were expressed as percentage of total cases. Study of significance was analysed by Chi-square test for qualitative data and Student T test for quantitative data. P value <0.05 was considered significant. Statistica version 6 [Tulsa, Oklahoma: StatSoft Inc., 2001] was used for statistical analysis. Numerical variables in the descriptive statistics were found to be normally distributed by Kolmogorov-Smirnov goodness-of-fit test., other than age of 1^st^ transfusion, age of diagnosis, liver enlargement. Genotypes were divided into following 3 groups for statistical analysis: Group 1-1-5/1-5; Group 2-1-5/HbE; Group 3-1-5/None of 5. Comparison of numerical variables between Groups 1, 2 and 3 were done by One-way Analysis of variance (ANOVA) and Kruskal-Wallis ANOVA.

## Results

In the study population of 103 subjects, 61 (59.22%) were male and 42 (40.78%) were female; 74 (71.48%) were TDT and 29 (28.16%) were NTDT. The distribution of subjects as per HPLC analysis is presented in supplementary table 1. Fifty-four patients (52.43%) were beta major, 3 patients (2.91%) were beta intermedia, 45 patients (43.69%) were E beta and 1 patient (0.97%) was S beta. Representative multiplex touchdown ARMS PCR gel image is shown in figure 1. The proportion of different genotypes and alleles detected is presented in supplementary table 2 and supplementary table 3 respectively. Among 63 TDT patients, 35 patients (55.56%) had 1-5/1-5 mutation, 15 (23.81%) had 1-5/HbE and 13 (20.63%) had 1-5/None of 5 mutation and among 23 NTDT patients, 20 patients (86.96%) had 1-5/HbE mutation and 3 (13.04%) had 1-5/1-5 mutation. The mean age of starting transfusion, mean age of diagnosis, mean rate of blood transfusion, mean hemoglobin level at diagnosis among different genotypes are presented in figure 2. Patients belonging to the three different genotype groups significantly differed in all these parameters except for mean hemoglobin level at presentation. Hb HPLC findings were discordant in 23 cases (22.55%). Diagnosis of 23 (22.55%) patients were not feasible by Hb-HPLC but confirmed by genetic mutation study. We also identified several rare pathogenic mutations by sequencing the *HBB* gene like c.316-106C>G (dbSNP: 34690599); Hb Kairouan (c.92G>C); c.33C>A (dbSNP rs35799536); c.47G>A (dbSNP rs63750783); c.51delC (HbVar ID 799); c.[93-2A>C] and c.118C>T (HbVar ID 845) (figure 3). We also detected a novel Pathogenic M_000518.5(*HBB*):c.164_168delinsGGCATCA (p.Val55fs) mutation in heterozygous state which was reported in the ClinVar database with accession ID VCV000590977.2 (figure 4).

**Table 1.**
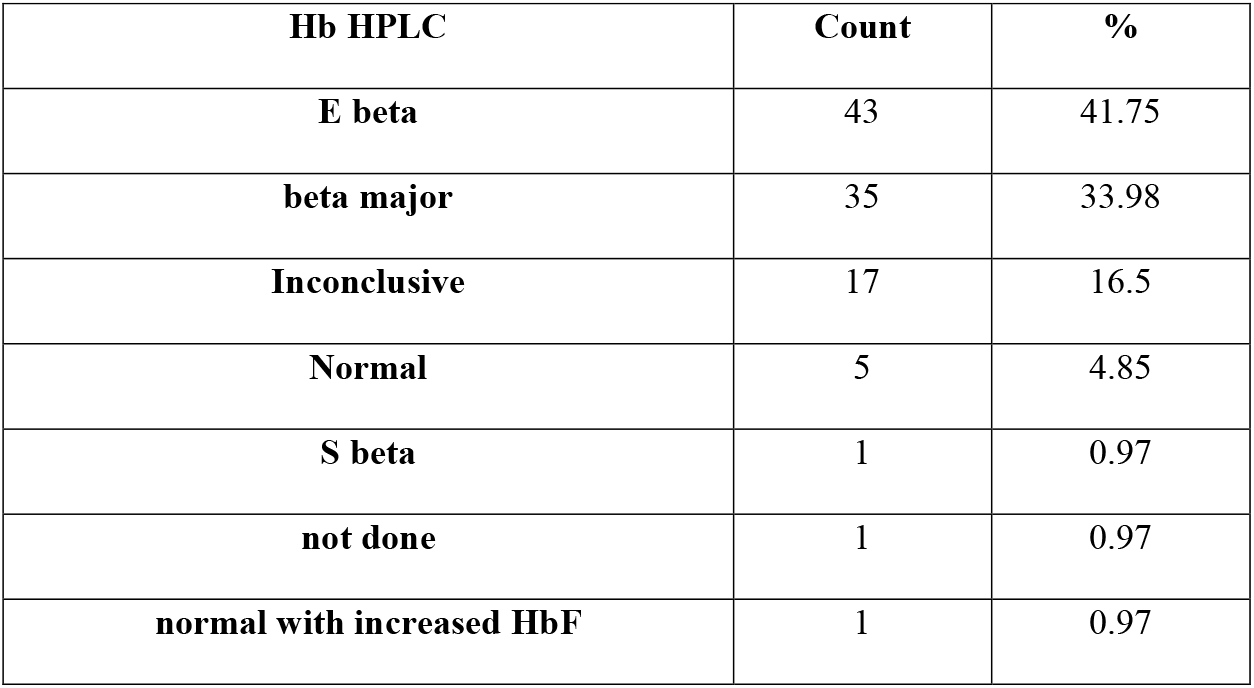
Distribution of patients according to haemoglobin HPLC report.

**Table 2.**
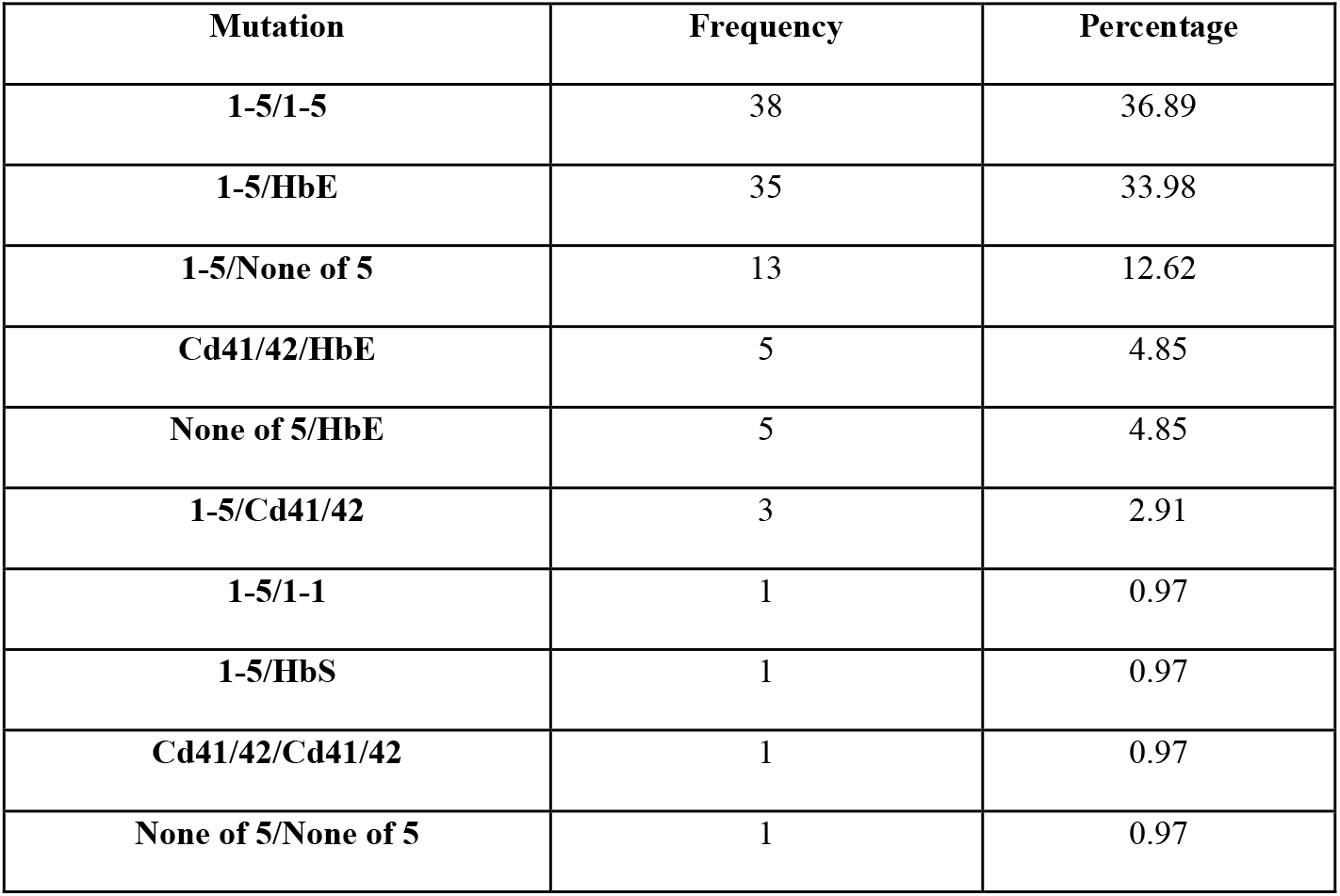
Proportion of different genotypes.

**Table 3.**
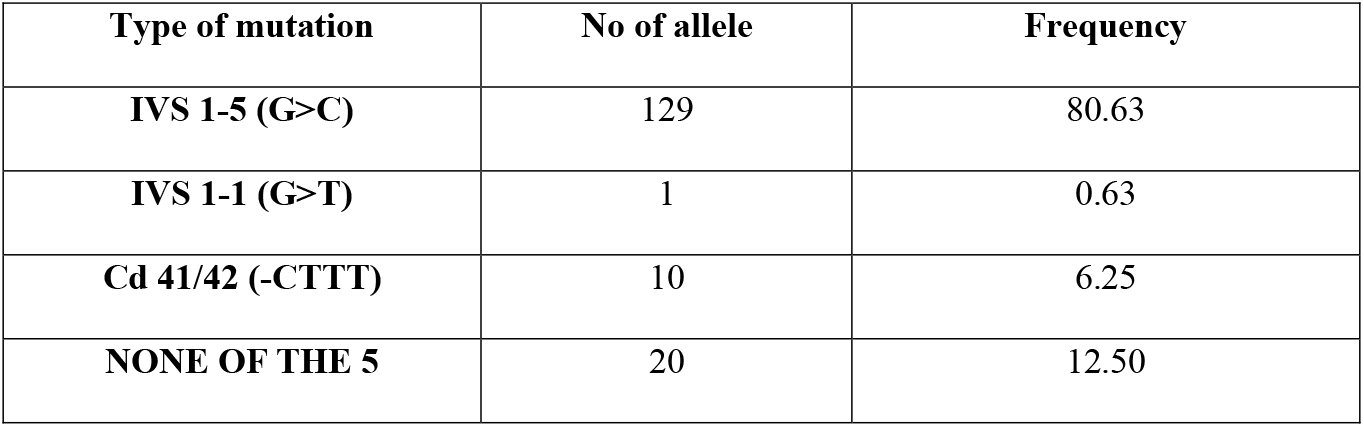
Distribution of mutant alleles.

**Figure 1.**
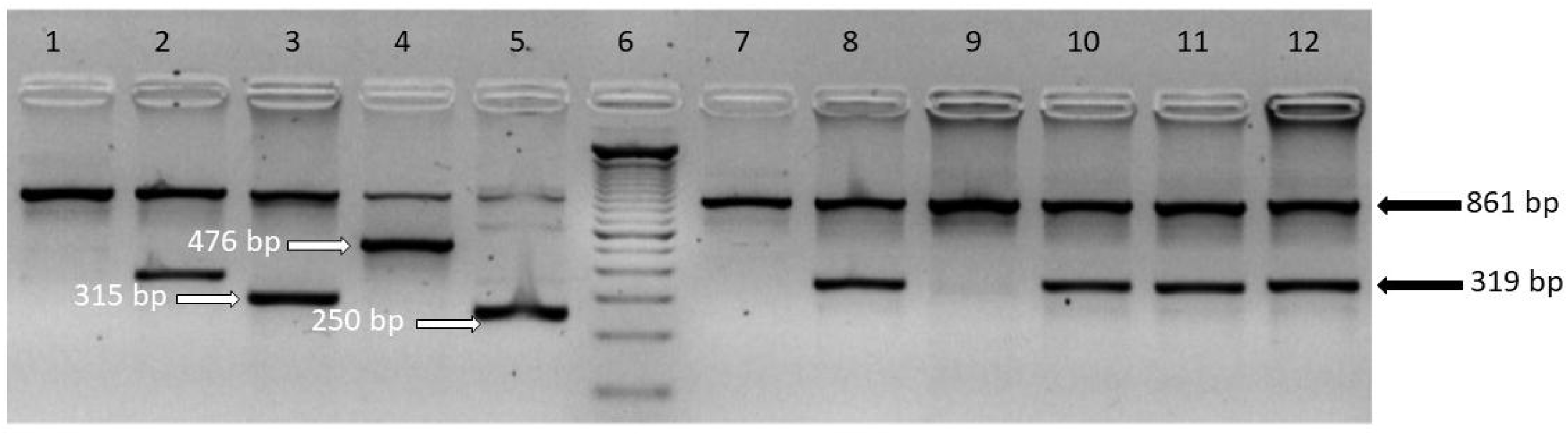
Representative gel image of multiplex ARMS PCR for β Thalassaemia five common mutations. 861 bp band in all the wells denote the control band. Lane 6 demonstrates the DNA ladder starting from 100 bp in 100 bp increments. Lane 1, 7 & 9 demonstrate absence of the five common mutations. Lane 2, 8, 10, 11 & 12 demonstrate the commonest IVSI-5 mutation denoted by the 319 bp mutant allele band. Lane 3 demonstrate IVS 1-1 mutation denoted by the 315 bp mutant allele band. Lane 5 demonstrate Cd 8/9 (+G) mutation denoted by the 250 bp mutant allele band. Lane 4 demonstrate Cd 41/42 mutation denoted by the 476 bp mutant allele band.

**Figure 2.**
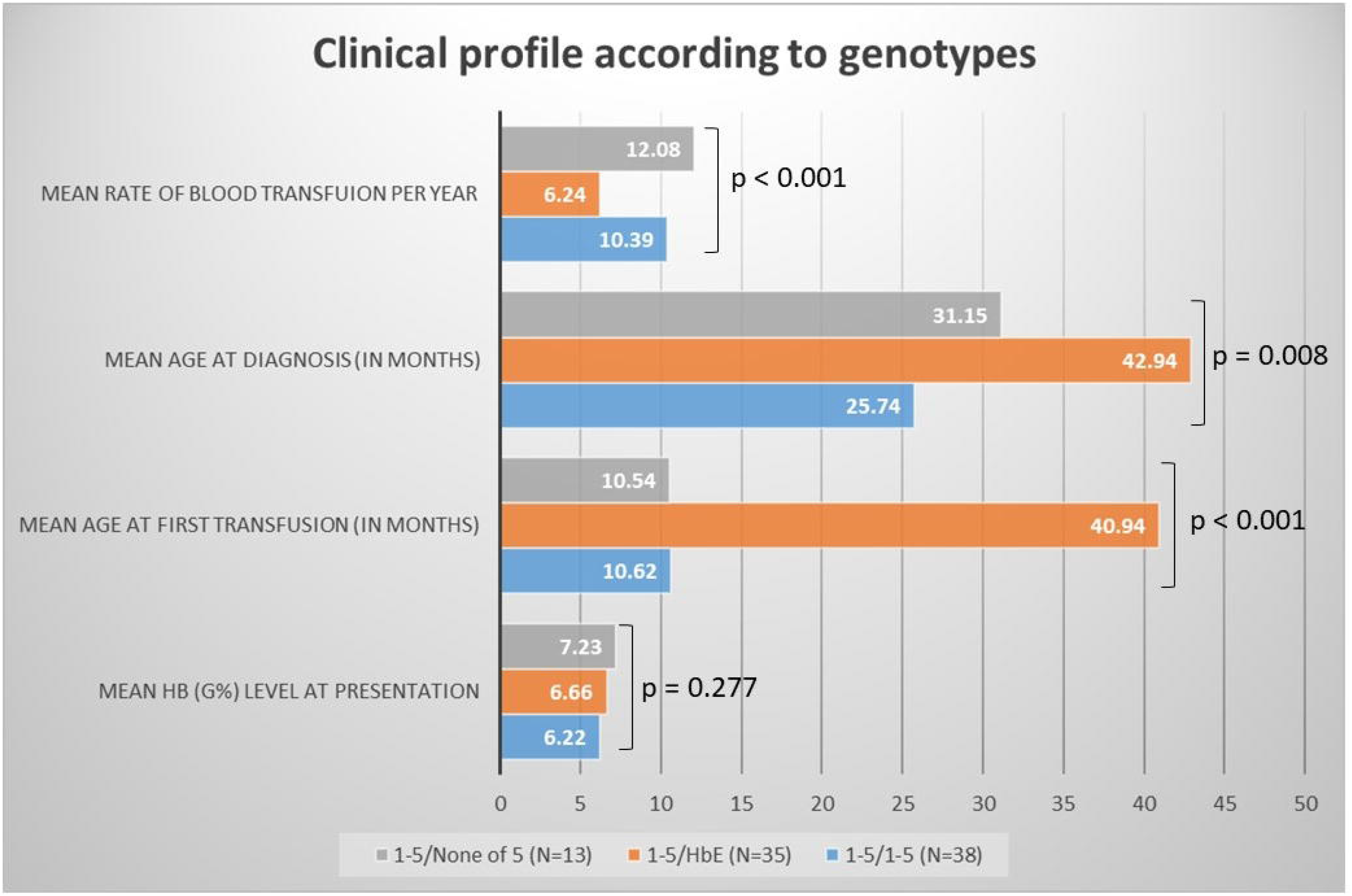
Bar diagram comparing the clinical profiles of the subjects classified according to their genotypes

**Figure 3.**
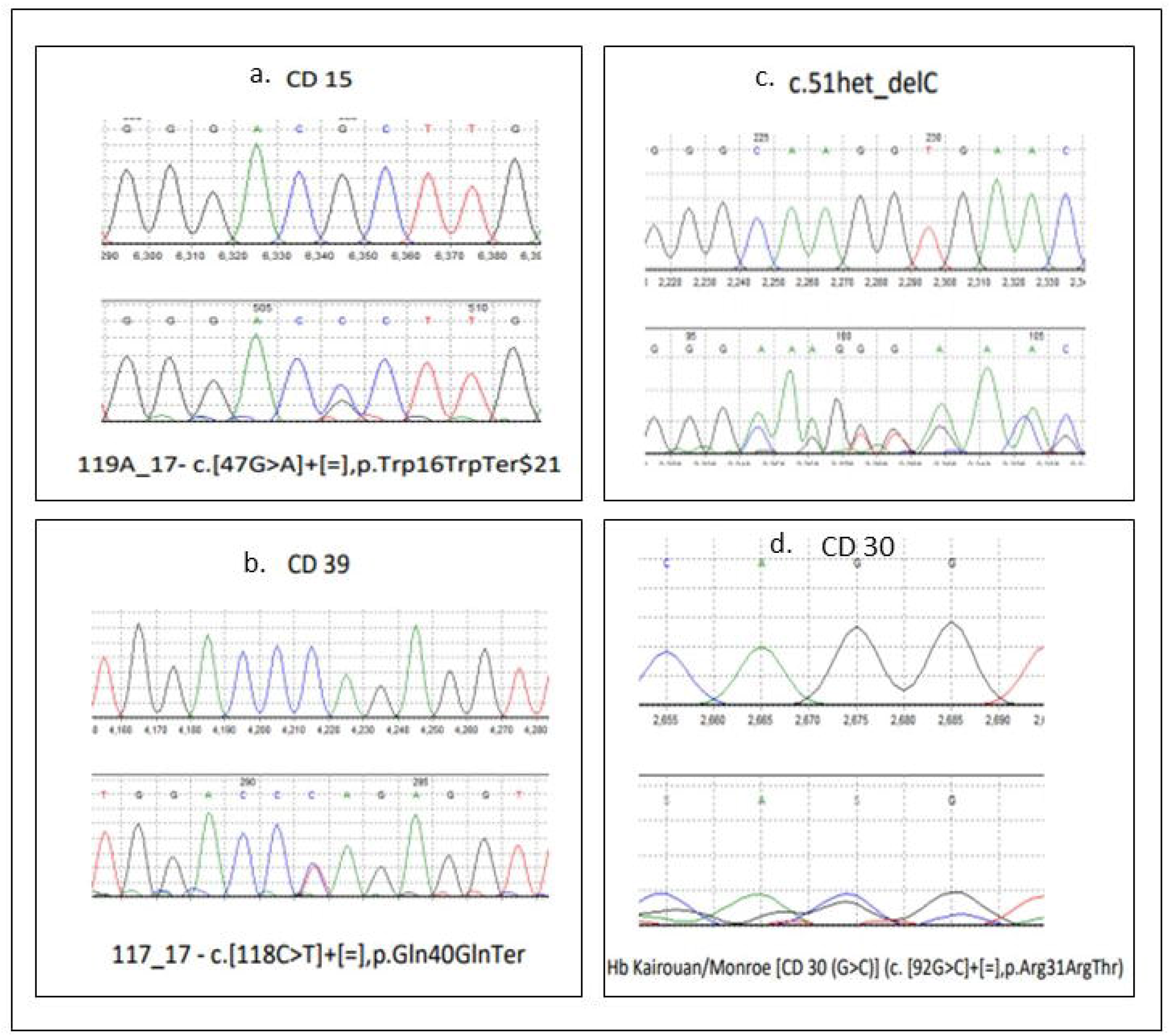
Capillary electrophoresis diagram of four rare pathogenic variants in the *HBB* gene. The top panel represent wild type and bottom panel the mutant allele for each mutation.

**Figure 4.**
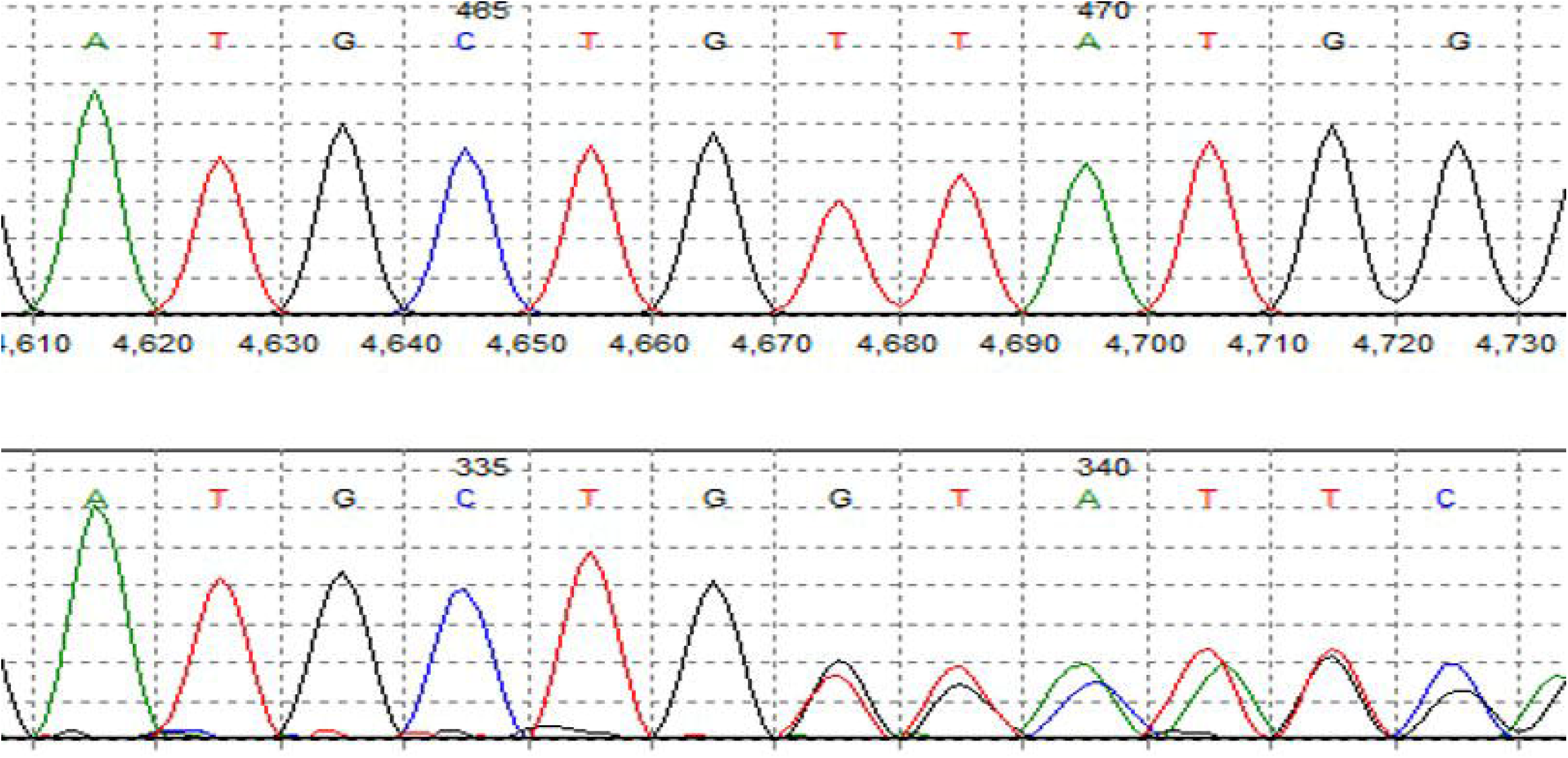
Capillary electrophoresis diagram of the novel Pathogenic M_000518.5(*HBB*):c.164_168delinsGGCATCA (p.Val55fs) mutation in heterozygous state which was reported in the ClinVar database with accession ID VCV000590977.2.

## Discussion

The present study revealed the distribution of the *HBB* gene mutations causing β Thalassemia along with a phenotype-genotype correlation in a cohort (n=103) of paediatric population in West Bengal. In this study among the β/β Thalassemia patients, 54 were TDT i.e., β major and 3 were NTDT i.e., β intermedia. Out of 54 TDT patients, 38 (70.37%) had IVS 1-5/1-5 mutation. Therefore, strong correlation of severity of phenotype and genotype was found in β Thalassemia symptomatic children with IVS 1-5 mutation, which was the commonest mutation found in this cohort. Out of 45 E β Thalassemia patients, 20 were TDT and 25 were NTDT. So, same 1-5 mutation when combined with HbE then phenotypic severity was modified by some unknown modifier as revealed in this study. So, genotype-phenotype correlation was not found in E β Thalassemia with commonest genotype (1-5/HbE) in this cohort. HPLC screening for carriers especially in the antenatal setting is well established in the health system. It is also the primary investigation in Thalassemia cases. In this study, HPLC findings were discordant in23/102 cases i.e. 22.55% (95% Confidence Interval 14.44 to 30.66%). Those patients were ultimately diagnosed by genetic mutation study. Further, 2 silent carrier fathers were diagnosed by genetic study, i.e.,1-5 heterozygous. It was clear, genetic mutation study confirmed the diagnosis where HPLC was inconclusive. In this study, it was highlighted that there was a definite delay in the mean age of diagnosis (in 1-5/1-5 genotype-10.62±10.459 months) from the mean age of 1st blood transfusion (in same genotype-25.74±20.105 months). This was as because those patients presented very early within first year in remote areas and started getting blood transfusion every month. Genetic mutation study can fill this gap, even confirm the diagnosis in antenatal period. Further study needs to define the genetic modification which interacts with β-Thalassemia, to provide better understanding of the process of the disease. Present study highlights only 2 mutations (IVS 1-5 and Cd41/42) plus HbE mutation study detected 79.6% of symptomatic β Thalassemia in study population unlike much higher detection rate with common five mutations study in other parts of India. In this study IVS 1-5 mutations evolved as most common mutation which was predictor of clinical severity in symptomatic β/β Thalassemia but not in E β Thalassemia.

In conclusion, the present study corroborated that most common type of mutation of β allele in this eastern part of India is IVS 1-5 (G>C) and that genotype and phenotype correlation was found in β/β Thalassemia but not in E β Thalassemia. In addition, we detected many rare variants including a novel indel variant (VCV000590977.2) in the *HBB* gene. This study emphasized the role of genetic mutation analysis as an adjunct to hemoglobin HPLC.

## Data Availability

All data produced in the present work are contained in the manuscript

## Ethical Approval

Institutional ethics committee had approved the current work.

## Informed Consent

All children were included in this study after obtaining written informed consent from their legal guardians.

## Consent for publication

All children were included in this study after obtaining written informed consent from their legal guardians regarding publication of the findings of the study without disclosing their identity.

## Acknowledgments

Professor (Dr.) Avijit Hazra, Dept of Pharmacology for helping with the statistical analysis Patients and their Parents for their continuous co-operation.

