## Supplementary data for "The spectrum of rare and novel indel mutations responsible for β Thalassemia in eastern India"

**Supplementary protocol 1**

Molecular assay for β Thalassemia five common mutations of Asian Indians by Multiplex ARMS PCR

### Summary:

The protocol described here is a modified form of the method described by Bhardwaj *et al*, 2005 for five common mutations accounting for ~ 90% cases of β Thalassemia in Asian-Indians.

### Introduction:

β Thalassemia (OMIM 613985) is an autosomal recessive haemolytic anaemia with a reported prevalence of ~8% in West Bengal (β-thalassemia trait 6.61%, β- thalassemia major 0.73%, compound heterozygosity for HbE-β-thalassemia 0.42%)^1^. β Thalassemia can be caused by homozygous or compound heterozygous mutation in the beta-globin gene (HBB) on chromosome 11p15. Beta-thalassemia is characterized by a reduced production of hemoglobin A (HbA, alpha-2/beta-2), which results from the reduced synthesis of beta-globin chains relative to alpha- globin chains, thus causing an imbalance in globin chain production and hence abnormal erythropoiesis. The disorder is clinically heterogeneous .Absence of beta globin causes beta-zero-thalassemia. Reduced amounts of detectable beta globin causes beta-plus-thalassemia. For clinical purposes, beta-thalassemia is divided into thalassemia major (transfusion dependent), thalassemia intermedia (of intermediate severity), and thalassemia minor (asymptomatic, carrier state). The remarkable phenotypic diversity of the beta-thalassemias reflects the heterogeneity of mutations at the HBB locus, the action of many secondary and tertiary modifiers, and a wide range of environmental factors.

ARMS PCR:

Amplification refractory mutation system has been used as a robust mutation detection method for many inherited disorders. The current protocol describes a modified version of multiplex ARMS PCR method originnay described by Bhardwaj *et al,* 2005 which detects IVSI-5 (G-C), IVSI-1 (G-T), Cd 8/9 (+G), Cd 41/42 (-TTCT) and 619 bp deletion.

### Basic protocol 1:

**Materials:**

a) PCR mix (200 μM dNTPs, 1.5 mM MgCl2, 10X buffer)
b) Primers
c) MiliQ water
d) Taq DNA polymerase
e) Control and test samples
f) 1X TBE buffer
g) Agarose powder
h) 6X Gel loading dye – bromophenol blue
i) Ethidium bromide
j) ABI Veriti 96 well thermal cycler
k) Horizontal submarine electrophoresis system with power pack
l) Biorad Gel documentation system.

### Notes:

I L 5X Tris-EDTA buffer contains

a. 54 g of Tris base
b. 27.5 g of boric acid
c. 20 mL of 0.5 M EDTA (pH 8.0)

### DNA extraction:

Refer to SOP of DNA extraction from venous blood

### Primers used:

| Sl no | Name | Sequence (5`-3`) | Tm (^0^C) | PCR  produ ct size |
| --- | --- | --- | --- | --- |
| 1 | Thal IVSI-5 (G-C) (mutant) | CTC CTT AAA CCT GTC TTG TAA CCT TGT TAG | 64 | 319 |
| 2 | Thal IVSI-5 (G-C) (wild) | CTC CTT AAA CCT GTC TTG TAA CCT TGT TAC |  |  |
| 3 | Thal IVSI-1 (G-T) (mutant) | TTA AAC CTG TCT TGT AAC CTT GAT ACG AAA | 61.3 | 315 |
| 4 | Thal IVSI-1 (G-T) (wild) | GAT GAA GTT GGT GGT GAG GCC CTG GGT AGG |  |  |
| 5 | Thal Cd 8/9 (+G) (mutant) | CCT TGC CCC ACA CGG CAG TAA CGG CAC ACC | 75 | 250 |
| 6 | Thal Cd 8/9 (+G) (wild) | CCT TGC CCC ACA GGG CAG TAA CGG CAC ACT |  |  |
| 7 | Thal Cd 41/42 (mutant) | GAG TGG ACA GAT CCC CAA AGG ACT CAA CCT | 69.5 | 476 |
| 8 | Thal Cd 41/42 (wild) | GAG TGG ACA GAT CCC CAA AGG ACT CAA AGA |  |  |
| 9 | Thal common E | TGA AGT CCA ACT CCT AAG CCA GTG | 62.7 | - |
| 10 | Thal control A | CAA TGT ATC ATG CCT CTT TGC ACC | 61 | 861 |
| 11 | Thal control B | GAG TCA AGG CTG AGA GAT GCA GGA | 64.4 |  |

To make the working primer solution each primer stock is 10 fold diluted in Mili Q water to make a final volume of 50 μL (5 μL stock + 45 μL Mili Q water). The stock and working primer solutions are kept in -20^0^ C refrigerator until used.

### PCR protocol:

**PCR mix for X number of samples (if X is less or equal to 20 make mix for X+1 and if more then X+2). Final volume 25.05 μL. Three separate tubes** for NTC and positive and negative controls to be used along with test samples.

| Component | Volume (in **μL)** |
| --- | --- |
| PCR mix ( 200 μM dNTPs, 1.5 mM MgCl2, 10X buffer) | 4.2 x (X+1) |
| 7 primers (0.75 **μL each)** | 5.25 x (X+1) |
| Taq DNA polymerase | 0.25 x (X+1) |
| Water | 13.35 x (X+1) |
| DNA (water for NTC) | 2 x (X+1) |
| Total volume | 25.05 x (X+1) |

**PCR conditions: Touchdown PCR (58^0^ C to 65^0^ C) Initial 14 cycles:**

| Cycle name | Temperature in ^0^C | Time |
| --- | --- | --- |
| Initial denaturation | 95 | 10 minutes |
| Denaturation | 94 | 45 seconds |
| Annealing | 65 | 60 seconds |
| Extension | 72 | 60 seconds |

Next 25 cycles:

| Cycle name | Temperature in ^0^C | Time |
| --- | --- | --- |
| Denaturation | 94 | 45 seconds |
| Annealing | 58 | 60 seconds |
| Extension | 72 | 60 seconds |
| Final extension | 72 | 7 minutes |
| Hold | 4 | Till tubes are taken out |

PCR tubes are kept at 4^0^ C until electrophoresis.

### Interpretation:

Prior to interpretation the carrier / affected status of the subjects tested based on hemoglobin electrophoresis data should be known to the laboratory in-charge. Absence of any band could either represent PCR failure or homozygous 619 bp deletion. Presence of control band and absence of any other band rules out five common mutations. Presence of a single band in addition to control band can be present in either a carrier or a mutant homozygote. Presence of two bands in addition to control band denotes compound heterozygosity.

### Literature Cited:

### 1. Bhardwaj U, Zhang YH, Lorey F, McCabe LL, McCabe ERB. Molecular Genetic Confirmatory Testing from Newborn Screening Samples for the Common African- American, Asian Indian, Southeast Asian, and Chinese ß – Thalassemia Mutations. American Journal of Hematology 78: 249- 255 (2005).

**Supplementary protocol 2**

**Molecular analysis for HbE disease by PCR -RFLP**

### Summary:

The protocol described here is a modified form of the method described by Tachavanich *et al*, 2009 for m**olecular analysis for HbE disease by PCR -RFLP** .

### Introduction:

Haemoglobin E disease is a variant Hemoglobin with a mutation in the ß globin gene causing substitution of glutamic acid for lysine at position 26 of the ß globin gene. HbE disease is common in Southeast Asia with a prevalence of 3.92% (HbE trait) in Kolkata, West Benagl and 23.9% in Dibrugarh, Assam (Mohanty *et al*, 2013). The β chain of HbE (βE) is synthesized at a reduced rate compared with that of normal adult hemoglobin (HbA), as the mutation creates an alternate splicing site within an exon. Consequently heterozygotes AE, compound heterozygotes SE and homozygotes EE show some β thalassemic features. Subjects heterozygous for HbE (AE) have an asymptomatic condition with no clinical relevance, except for the risk of transmitting E/β thalassemia if the other parent carries β thalassemia.

**Basic protocol 1:**

**Materials:**

a) PCR mix (200 μM dNTPs, 1.5 mM MgCl2, 10X buffer)
b) Primers
c) MiliQ water
d) Taq DNA polymerase
e) Control and test samples
f) MnlI restriction enzyme
g) NEB Buffer.
h) 1X TBE buffer
i) 100 bp DNA ladder
j) Agarose powder
k) 6X Gel loading dye – bromophenol blue
l) Ethidium bromide
m) ABI Veriti 96 well thermal cycler
n) Horizontal submarine electrophoresis system with power pack
o) Biorad Gel documentation system.
p) Water bath.

**Primers and reconstitution procedure:**

| Sl no | Name | Sequence (5`-3`) | Tm (^0^C) | ΜL Mili Q water added to make 100pmol | PCR  produ ct size |
| --- | --- | --- | --- | --- | --- |
| 1 | HbE-F | CAT TTG CTT CTG ACA CAA CTG | 55.9 | 501 | 427 |
| 2 | HbE-R | TTG AGG TTG TCC AGG TGA | 56.7 | 462 | 427 |

To make the working primer solution each primer stock is 10 fold diluted in Mili Q water to make a final volume of 50 μL (5 μL stock + 45 μL Mili Q water). The stock and working primer solutions are kept in -20^0^ C refrigerator until used.

### PCR protocol:

**PCR mix for X number of samples (if X is less or equal to 20 make mix for X+1 and if more then X+2). Final volume 20 μL. Three separate tubes for NTC and positive and negative controls to be used along with test samples.**

| Component | Volume (in **μL)** |
| --- | --- |
| PCR mix ( 100 μM dNTPs, 1.5 mM MgCl2, 10X buffer) | 3.36 x (X+1) |
| 2 primers (0.60 **μL each)** | 1.20 x (X+1) |
| Taq DNA polymerase | 0.20 x (X+1) |
| Water | 13.24 x (X+1) |
| DNA (water for NTC) | 2 x (X+1) |
| Total volume | 25.00 x (X+1) |

### PCR conditions: Touchdown PCR (56^0^ C to 63^0^ C) Initial 14 cycles:

| Cycle name | Temperature in ^0^C | Time |
| --- | --- | --- |
| Initial denaturation | 95 | 10 minutes |
| Denaturation | 94 | 45 seconds |
| Annealing | 63 | 60 seconds |
| Extension | 72 | 60 seconds |

Next 25 cycles:

| Cycle name | Temperature in ^0^C | Time |
| --- | --- | --- |
| Denaturation | 94 | 45 seconds |
| Annealing | 56 | 60 seconds |
| Extension | 72 | 60 seconds |
| Final extension | 72 | 7 minutes |
| Hold | 4 | Till tubes are taken out |

PCR tubes are kept at 4^0^ C until electrophoresis.

### Restriction digestion with MnlI:

| Component | Volume (in **μL)** |
| --- | --- |
| Cutsmart buffer (10X) | 1.50x (X) |
| **MnlI (5 U)** | 1.00 x (X) |
| PCR product | 5.00 x (X) |
| Water | 7.50 x (X) |
| Total volume | 15.00 x (X) |

Incubate at 37ºC for 1 hour in air bath.

### Interpretation:

HbE normal homozygote

| # | Ends | Coordinates | Length |
| --- | --- | --- | --- |
| 1 | MnlI-MnlI | 122-292 | 171 |
| 2 | MnlI- (Right End) | 293-427 | 135 |
| 3 | MnlI-MnlI | 62-121 | 60 |
| 4 | (LeftEnd)- MnlI | 1-45 | 45 |
| 5 | MnlI-MnlI | 46-61 | 16 |

HbE mutant homozygote

| # | Ends | Coordinates | Length |
| --- | --- | --- | --- |
| 1 | MnlI-MnlI | 62-292 | 231 |
| 2 | MnlI- (Right End) | 293-427 | 135 |
| 3 | (LeftEnd)- MnlI | 1-45 | 45 |
| 4 | MnlI-MnlI | 46-61 | 16 |

### HbE trait:

| # | Ends | Coordinates | Length |
| --- | --- | --- | --- |
| 1 | MnlI-MnlI | 62-292 | 231 |
| 2 | MnlI-MnlI | 122-292 | 171 |
| 3 | MnlI- (Right End) | 293-427 | 135 |
| 4 | MnlI-MnlI | 62-121 | 60 |
| 5 | (LeftEnd)- MnlI | 1-45 | 45 |
| 6 | MnlI-MnlI | 46-61 | 16 |

**Supplementary table 1**

**Distribution of patients according to haemoglobin HPLC report**

| **Hb HPLC** | **Count** | **%** |
| --- | --- | --- |
| **E beta** | 43 | 41.75 |
| **beta major** | 35 | 33.98 |
| **Inconclusive** | 17 | 16.5 |
| **Normal** | 5 | 4.85 |
| **S beta** | 1 | 0.97 |
| **not done** | 1 | 0.97 |
| **normal with increased HbF** | 1 | 0.97 |

**Supplementary table 2**

**Proportion of different genotypes**

| **Mutation** | **Frequency** | **Percentage** |
| --- | --- | --- |
| **1-5/1-5** | 38 | 36.89 |
| **1-5/HbE** | 35 | 33.98 |
| **1-5/None of 5** | 13 | 12.62 |
| **Cd41/42/HbE** | 5 | 4.85 |
| **None of 5/HbE** | 5 | 4.85 |
| **1-5/Cd41/42** | 3 | 2.91 |
| **1-5/1-1** | 1 | 0.97 |
| **1-5/HbS** | 1 | 0.97 |
| **Cd41/42/Cd41/42** | 1 | 0.97 |
| **None of 5/None of 5** | 1 | 0.97 |

**Supplementary table 3**

**Distribution of mutant alleles**

| **Type of mutation** | **No of allele** | **Frequency** |
| --- | --- | --- |
| **IVS 1-5 (G>C)** | 129 | 80.63 |
| **IVS 1-1 (G>T)** | 1 | 0.63 |
| **Cd 41/42 (-CTTT)** | 10 | 6.25 |
| **NONE OF THE 5** | 20 | 12.50 |
